# Physician Approaches to the Pharmacologic Treatment of Dystonia in Cerebral Palsy

**DOI:** 10.1101/2024.02.01.24302121

**Authors:** Emma Lott, Darcy Fehlings, Rose Gelineau-Morel, Michael Kruer, Jonathan W. Mink, Sruthi P. Thomas, Steve Wisniewski, Bhooma Aravamuthan, the Cerebral Palsy Research Network

## Abstract

**Objective:** To determine how physicians approach pharmacologic dystonia treatment in people with CP and assess physician readiness to participate in a randomized trial comparing existing pharmacologic dystonia treatments.

**Methods:** We administered a REDCap survey to physician members of the American Academy of Cerebral Palsy and Developmental Medicine and of the Child Neurology Society to assess which pharmacologic agents they use to treat dystonia in CP and their preferred indications and dosing.

**Results:** Of 479 physicians surveyed, 240 (50%) responded. Respondents treated functionally limiting (95%) and generalized (57%) dystonia and most commonly used six medications: baclofen (95%), trihexyphenidyl (79%), gabapentin (67%), carbidopa/levodopa (55%), clonazepam (55%), and diazepam (54%). Baclofen was preferred in people with co-existing spasticity (81%), gabapentin was preferred in people with co-existing pain (49%), and trihexyphenidyl was avoided in people with constipation (34%) or urinary retention (42%). Preferred dosing regimens followed published regimens for dystonia, when available, but otherwise followed published regimens for other CP symptoms (spasticity and seizures). Baclofen was preferred by 64% of respondents as first line treatment, but there was no clear consensus on second or third-line medications. Most respondents (51%) were comfortable randomizing their patients to receive any of the six most commonly used medications used to treat dystonia in CP.

**Conclusions:** This study summarizes current indications and dosing for the six most commonly used medications to treat dystonia in CP as per treating physicians in the US and Canada and also demonstrates physician support for a randomized trial comparing the effectiveness of these treatments.

**Article summary:** Dystonia is common and debilitating in people with CP, with little data on pharmacologic treatments. We describe physicians’ current approaches to using these treatments.

**What’s known on this subject:** Comparing the effectiveness of existing pharmacologic treatments for dystonia in CP is a research priority shared by clinicians and the community. However, current pharmacologic treatment practices are unknown.

**What this study adds:** Physicians in the US and Canada primarily prescribe a subset of six medications for the treatment of functionally limiting and generalized dystonia in CP: baclofen, trihexyphenidyl, gabapentin, carbidopa/levodopa, clonazepam, and diazepam.

**Contributors Statement:** Emma Lott helped design the study, carried out data analyses, and critically reviewed and revised the manuscript.

Darcy Fehlings, Rose Gelineau-Morel, Michael Kruer, Jonathan Mink, Sruthi Thomas, and Steve Wisniewski helped design the study and critically reviewed and revised the manuscript.

Bhooma Aravamuthan conceptualized and designed the study, supervised data collection and analysis, drafted the initial manuscript, and critically reviewed and revised the manuscript.

## Introduction

Cerebral palsy (CP) is the most common childhood-onset motor disability and the most common condition associated with dystonia in young people.^1–3^ Dystonia is an often painful and debilitating movement disorder characterized by overflow muscle activation triggered by attempted voluntary movement.^4–6^ Up to 80% of people with CP are affected by dystonia.^7^ Though multiple pharmacologic agents are available to treat dystonia in people with CP, it is unclear which, if any, are effective.^8^ Therefore, it is necessary to compare the effectiveness of existing treatments, a top research priority that emerged in a recent collaborative effort between clinicians, researchers, and the community to identify areas of research need for dystonia in CP.^9^

Current guidance for treatment of dystonia in people with CP is based largely on expert opinion. The American Academy of Cerebral Palsy and Developmental Medicine (AACPDM) Care Pathway for Dystonia in Cerebral Palsy recommends baclofen first line and trihexyphenidyl second line as pharmacologic treatment.^10^ Specific indications for other medications included consideration of gabapentin for people with dystonia and pain and clonidine for people with dystonia and poor sleep.^10^ The data supporting these recommendations are of low certainty.^8^

To compare the effectiveness of existing pharmacologic treatments for dystonia in CP, we must first establish how pharmacologic treatments are currently used. In this study, we queried how physicians in the US and Canada approach pharmacologic dystonia treatment in people with CP and assessed physician readiness to participate in a randomized trial comparing the efficacy of these medications. We hypothesized that physicians would have variable treatment approaches and would generally support a trial comparing their efficacy to treat dystonia in CP.

## Methods

### Standard Protocol Approvals, Registrations, and Patient Consents

Human Subjects Research exemption was granted by the Washington University Institutional Review Board (IRB ID# 201910233, 11/04/2019).

### Surveyed population

We surveyed the physician memberships of the AACPDM and select Special Interest Groups (SIGs) within the Child Neurology Society (CNS): the CP SIG and the Movement Disorders SIG. Physicians primarily practicing outside the US or Canada were excluded. Medical specialty data of non-responders was abstracted through society membership data.

### Survey development and administration

The survey was developed in REDCap via iterative discussions between medical specialists in child neurology (BRA, RGM, JWM), developmental pediatrics (DF), physiatry (ST), and an epidemiologist and clinical trialist (SW). Respondents’ approaches to using the following 10 medications were queried explicitly with the opportunity to write in other medications: baclofen, trihexyphenidyl, gabapentin, carbidopa/levodopa, clonazepam, diazepam, clonidine, tetrabenazine, clobazam, and cannabidiol. Question formats included multiple choice, checkbox, and open-ended responses (see Supplementary Methods for Survey PDF). The survey was emailed to potential respondents weekly between 5/31/2023 and 7/19/2023.

### Qualitative analysis

Open-ended responses were analyzed using a conventional content analysis approach.^11^ Two investigators coded all responses and resolved discrepancies via discussion.

### Data availability

Anonymized data will be shared by request from any qualified investigator.

## Results

### Respondent demographics

In total, 479 physicians were eligible to take the survey (360 from AACPDM plus 135 from the CNS Movement Disorders and CP SIGs minus 16 practicing outside the US or Canada). Of these 479 physicians emailed the survey, 240 responded (response rate of 50%) of whom 219 confirmed they currently cared for people with CP (91%). Most cared primarily for children (166/217, 76%) in an academic setting (172/218, 79%) in the US (196/218, 90%) independently for more than 5 years (165/218, 76%). Most respondents were physiatrists (83/216, 38%) or neurologists (66/216, 31%) with at least 25% of their clinical practices comprised of people with CP (142/214, 66%) (Table 1). Medical specialty representation was similar between survey respondents and non-respondents (Supplementary Table 1).

**Table 1.** Physician Respondent Demo graphics.

|  | Respondents who currently<br>care for people with CP<br>(N=219) | Respondents who currently<br>prescribe medications for<br>dystonia (N=154) |
| --- | --- | --- |
|  | n, % | n, % |
| Patient age |  |  |
| Children | 166, 76% | 117, 76% |
| Adults | 4, 2% | 4, 3% |
| Both | 47, 22% | 32, 21% |
| Total respondents | 217, 100% | 153, 100% |
| Practice setting |  |  |
| Academic | 172, 79% | 122, 79% |
| Private | 19, 9% | 13, 8% |
| Both | 27, 12% | 19, 12% |
| Total respondents | 218, 100% | 154, 100% |
| Practice location |  |  |
| US | 196, 90% | 138, 90% |
| Canada | 22, 10% | 16, 10% |
| Total respondents | 218, 100% | 154, 100% |
| Years in practice |  |  |
| In training | 9, 4% | 3, 2% |
| 0 to 5 | 44, 20% | 34, 22% |
| 6 to 10 | 47, 22% | 33, 21% |
| 11 to 15 | 28, 13% | 19, 12% |
| >15 | 90, 41% | 65, 42% |
| Total respondents | 218, 100% | 154, 100% |
| Specialty |  |  |
| Physiatry | 83, 38% | 74, 48% |
| Neurology | 66, 31% | 56, 37% |
| Orthopedics | 38, 18% | 5, 3% |
| Developmental Pediatrics | 13, 6% | 8, 5% |
| Neurodevelopmental<br>Disabilities | 6, 3% | 6, 4% |
| Complex Care Pediatrics | 3, 1% | 2, 1% |
| General Pediatrics | 2, 1% | 1, 1% |
| Other | 5, 2% | 1, 1% |
| Total respondents | 216, 100% | 153, 100% |
| % of patients with CP |  |  |
| <5 | 9, 4% | 6, 4% |
| 5 to 25 | 63, 29% | 42, 28% |
| 26 to 50 | 52, 24% | 32, 21% |
| 51 to 75 | 54, 25% | 40, 27% |
| >75 | 36, 17% | 30, 20% |
| Total respondents | 214, 100% | 150, 100% |

Most respondents relied on physical exam (210/219, 96%) and history (175/219, 80%) to diagnose dystonia in people with CP. Less common approaches included video review (39/219, 18%) or a validated tool or scale (46/219, 21%), most commonly the Hypertonia Assessment Tool (Table 2).

**Table 2.**
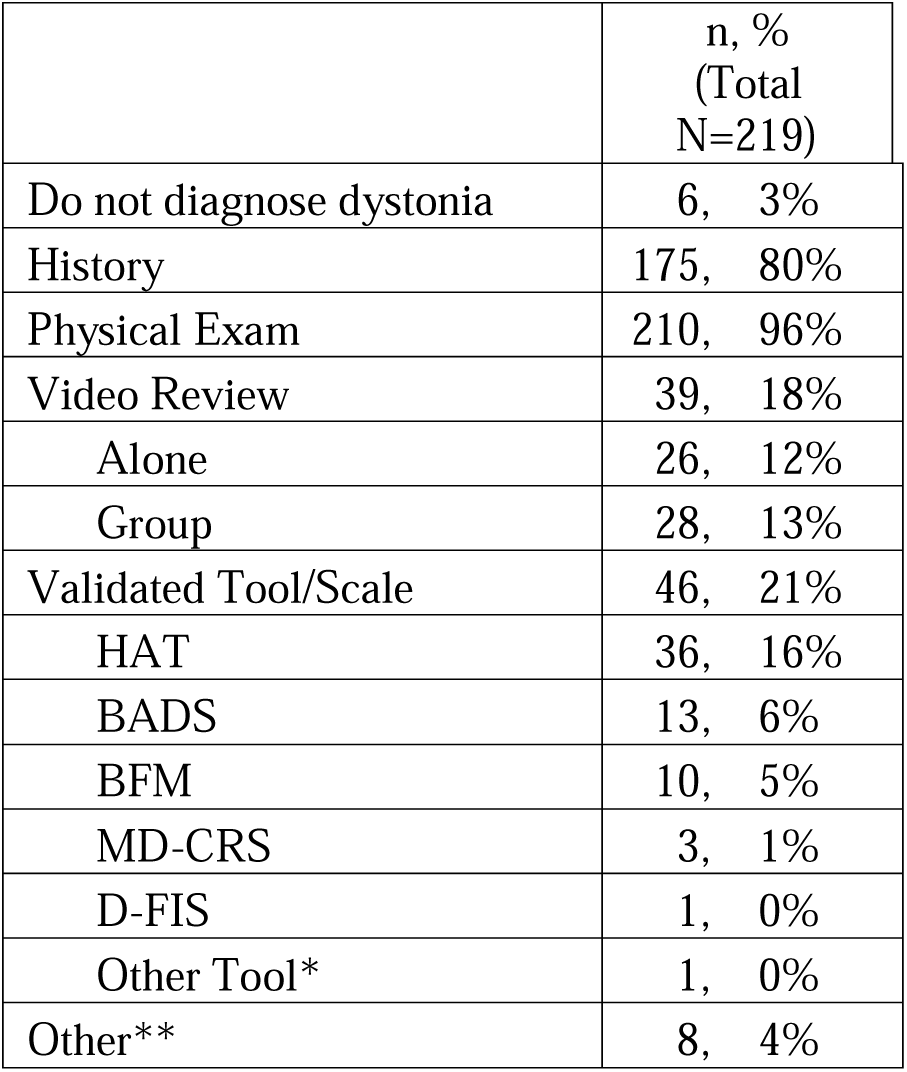
Methods used to diagnose dystonia. HAT – Hypertonia Assessment Tool, BADS – Barry Albright Dystonia Rating Scale, BFM – Burke-Fahn-Marsden Rating Scale, MD-CRS – Movement Disorders – Childhood Rating Scale, D-FIS – Dyskinetic Cerebral Palsy Functional Impact Scale. *Other tool – written in response for Unified Dystonia Rating Scale. **Other – respondents indicated they used other methods to diagnose dystonia but did not clarify further.

Most respondents prescribed medications (172/217, 79%), the majority of whom also confirmed that they prescribed medications to treat dystonia (154/172, 90%). These 154 physicians were asked to describe their prescribing practices further. The demographics of these physicians were comparable to those of the entire respondent group, except for a slightly higher representation of physiatrists (74/154, 48%) and neurologists (56/154, 37%) (Table 1).

### Factors affecting whether medications are prescribed at all to treat dystonia in people with CP

Over 50% of respondents prioritized at least one of three factors when deciding whether to prescribe medications for dystonia in people with CP: 1) dystonia severity (defined in the survey as “severity based on my assessment in the clinic”), 2) functional impact (defined as “whether a person’s dystonia prevents their ability to do a task that they deem is important to them, causes significant pain, interferes with sleep, or creates challenges associated with ease of caregiving”), and 3) whether the dystonia is focal or generalized (Table 3). Almost all physicians considered dystonia severity (144/154, 94%) and functional impact (146/154, 95%) when deciding whether to prescribe medications at all. Of the 61 people who provided written explanations for how they considered dystonia severity when deciding to prescribe medications, the majority (40/61, 66%) cited reasons having to do with functional impact:

> *“will prescribe depending on level of distress or dysfunction caused to the patient.”*

**Table 3.** Factors affecting whether to prescribe a medication and which medication to prescribe.

| Medical or demographic consideration | Affects whether to prescribe a medication at all | Affects the choice of which medication to prescribe |
| --- | --- | --- |
|  | n, %<br>(Total N=154) | n, %<br>(Total N=154) |
| Age | 55 36% | 89, 58% |
| Severity | 144 94% | 94, 61% |
| Focal versus Generalized | 101 66% | 103, 67% |
| Arms versus Legs | 44 29% | 44, 29% |
| Functional Impact | 146 95% | 95, 62% |
| GMFCS | 39 25% | 32, 21% |
| Etiology | 32 21% | 44, 29% |
| Medical Complexity | 58 38% | 84, 55% |
| Prevention of secondary MSK complications | 76 49% | 42, 27% |
| Peri-operative tone management | 61 40% | 41, 27% |
| Other | 8 5% | 23, 15% |

> *“Must have a life impact”*When elaborating on why functional impact affected their decision to prescribe medications, some respondents indicated that it was the primary driver of their decision to prescribe:

> *“Main deciding factor”*

> *“This is the key question I think”*The next most common factor governing respondents’ decision to prescribe medications was whether dystonia was focal vs. generalized (cited by 101/154, 66%). Of the 44 people who elaborated on how they applied this factor clinically, the majority (38/44, 86%) noted that they tended to prescribe enteral or “systemic” medications if dystonia was generalized, but preferred to use injectables like botulinum toxin if dystonia was focal.

Other factors were less commonly cited (by <50% of respondents). Interestingly, though etiology was relatively infrequently noted to affect the choice to prescribe medications (32/154, 21%), 6 respondents of the 12 who provided explanations stated that a genetic or suspected genetic etiology would make them more likely to prescribe a medication. Three more respondents suggested that basal ganglia injury would make them more likely to prescribe a medication.

> *“dopamine trial for all children with dystonia and CP to eval for [Dopa-responsive dystonia]”*

> *“May be more likely to try oral meds for dystonia if basal ganglia involvement”*Of the 23 respondents who explained why age was a factor affecting their decision to prescribe medications, 8 noted simply that they would not prescribe medications below a certain age, ranging from newborns to 2 years old. Eleven people noted that age limited their choice of medications. Interestingly, 3 people noted that dystonia could not yet be functionally impactful at young ages which is why they would not treat:

> *“I might be more careful with oral medications such as baclofen in very young children as they seem to experience more side effects “*

> *“age determines functional impairment “*

### Factors affecting choice of medication prescribed to treat dystonia in people with CP

The most common factor that respondents cited as governing their choice of medications (Table 3) was dystonia being focal vs. generalized (103/154, 67%). Almost all of those explaining further (28/30, 93%) reiterated that they would use injectables for focal dystonia and enteral medications for generalized dystonia. The next most cited factors were functional impact (95/154, 62%) and severity (94/154, 61%). Respondents factoring in severity (n=17) stated that greater severity dystonia would make them consider surgical interventions (4/17, 24%) or injectables together with enteral medications (3/17, 18%):

> *“consider [intrathecal baclofen pump] if generalized/severe and/or significant spasticity also, sometimes will do combo meds and injections if severe”*Pain management (6/17, 36%) and sleep (3/17, 18%) were prioritized by the 17 respondents explaining how they factor in functional impact:

> *“If sleep is issue - may reach for gabapentin or clonazepam as first line. If pain is issue, may use valium as first line.”*Age of the patient affected medication choice for 58% of respondents (89/154). Of the 32 respondents elaborating further, the most common single explanation was that age governed medication formulation or dose (7/32, 22%). When discussing specific medications, age was factored in most commonly for trihexyphenidyl (6/32, 19%), dopaminergic agents (5/32, 16%), and baclofen (4/32, 13%), but variably so:

> *“in littlest patients would not use [trihexyphenidyl] or dopa”*

> *“Anticholinergics are better tolerated in the young. Dopamine supplementation is more likely to be efficacious in the young, in my opinion.”*

> *“…I also avoid baclofen in those younger than 1.5 years old”*

> *“In a younger child I…have been using baclofen more with good response and tolerance*.Medical complexity (clarified in the survey as “e.g. need for G-tube, tracheostomy, or other specialty-based medical care”) was also cited as a factor governing medication choice (84/154, 55%). Respondents explaining further (n=29) avoided medications whose side effects might worsen co-existing conditions (13/29, 45%) and chose medications whose side effects may improve co-existing conditions (5/29, 17%):

> *“pick meds that might help with another concern as well (eg trihexyphenidyl if drooling is a big problem) or avoid worsening another condition (eg baclofen and seizure threshold)”*

> *“Example, if they have sialorrhea, I may use trihexyphenidyl for anticholinergic effects in addition to helping with dystonia…”*Concerns about polypharmacy (5/29, 17%) and available formulations (4/29, 14%) were also noted when prescribing enteral medications to people with high medical complexity:

> *“depends if other complex neurologic needs may not want to prescribe another medication in similar class”*

> *“if all [G-tube fed] sometimes I’ll start with meds easily available as liquid or dissolvable tabs, if lots of AEDs/benzos may be more/less likely to start with [clonazepam]”*Other factors were cited by less than a third of respondents. Notably, respondents who cited etiology as a factor affecting medication choice (44/154, 29%) went on to explain that genetic or idiopathic etiologies may prompt them to use carbidopa/levodopa (7/13, 54%):

> *“I do not find [carbidopa/levodopa] effective but if sometimes if the exam shows NO Spasticity and only dystonia, the etiology is unclear and the child has a normal brain MRI I may offer a quick [carbidopa/levodopa] trial”*

### Medication management

Almost all respondents gauged medication efficacy by asking the person with CP or their caregivers if there had been any improvement (150/154, 97%). The vast majority also gauged medication efficacy by establishing a shared functional goal before treatment and assessing progress toward achieving that goal (113/154, 73%), assessing changes in dystonia severity on physical exam (124/154, 81%), and side effect burden (124/154, 81%). A minority of respondents used validated dystonia scales (16/154, 10%), most commonly the Barry-Albright Dystonia Scale (7/16, 44%) or the Burke-Fahn-Marsden Scale (7/16, 44%).

If a medication was found to be ineffective, respondents were equally likely to add a second medication to the first medication (38/154, 25%), wean off the first medication before starting a second medication (37/154, 24%), or simultaneously wean the first medication while up-titrating the second medication (43/154, 28%). Thirty-three of 154 respondents (21%) indicated that they will utilize any of the above options depending on the specific patient situation.

### Most common medications used, indications, and dosing

The six most used medications were baclofen (147/154, 95%), trihexyphenidyl (122/154, 79%), gabapentin (103/154, 67%), carbidopa/levodopa (84/154, 55%), clonazepam (85/154, 55%), and diazepam (83/154, 54%). Overall, 80% of respondents (123/154) used a subset of only these six medications as their first through third-line pharmacologic treatments for dystonia in CP. Fifty-seven percent of respondents (88/154) used five of these six medications as their first through fifth-line treatment choices. These top six medications, including baclofen as the most commonly used medication, did not differ between neurologists and physiatrists or differ based on number of years in practice (Supplementary Table 3). Other medications (clonidine, tetrabenazine, clobazam, cannabidiol, and single write-in entries for amantadine and benztropine) were each used by less than 20% of all respondents to treat dystonia and are not described further due to the relative paucity of respondent data (Table 4).

**Table 4.** Medications used to treat dystonia in CP and frequency of use. Medications are listed by descending frequency of overall use. *Other – only 2 respondents wrote in medications they used that were different from the 10 medications explicitly queried: amantadine and benztropine.

|  | **At all** | **1st line** | **2nd line** | **3rd line** | **4th line** | **5th line** |
| --- | --- | --- | --- | --- | --- | --- |
| **Medications** | n, %<br>(Total N=154) | n, % | n, % | n, % | n, % | n, % |
| Baclofen | 147, 95% | 98, 64% | 18, 12% | 17, 11% | 10, 6% | 4, 3% |
| Trihexyphenidyl | 122, 79% | 21, 14% | 35, 23% | 31, 20% | 21, 14% | 14, 9% |
| Gabapentin | 103, 67% | 9, 6% | 25, 16% | 27, 18% | 19, 12% | 23, 15% |
| Clonazepam | 85, 55% | 11, 7% | 23, 15% | 20, 13% | 14, 9% | 17, 11% |
| Carbidopa/Levodopa | 84, 55% | 9, 6% | 18, 12% | 20, 13% | 20, 13% | 17, 11% |
| Diazepam | 83, 54% | 6, 4% | 23, 15% | 15, 10% | 25, 16% | 14, 9% |
| Clonidine | 30, 19% | 0, 0% | 7, 5% | 7, 5% | 8, 5% | 8, 5% |
| Tetrabenazine | 27, 18% | 0, 0% | 2, 1% | 6, 4% | 8, 5% | 11, 7% |
| Clobazam | 11, 7% | 0, 0% | 3, 2% | 2, 1% | 3, 2% | 3, 2% |
| Cannabidiol | 7, 5% | 0, 0% | 0, 0% | 3, 2% | 2, 1% | 2, 1% |
| Other* | 2, 1% | 0, 0% | 0, 0% | 2, 1% | 0, 0% | 0, 0% |

Most respondents used baclofen first-line to treat dystonia (98/154, 64%), but there was no clear consensus on the choice of 2^nd^ or 3^rd^ line medications. The indications and dosing regimens preferred by respondents for each of the top six medications used to treat dystonia in CP are shown in Table 5 and 6, respectively. Preferred dosing regimens for trihexyphenidyl^12–14^, gabapentin^15^, and carbidopa/levodopa^16^ were in line with published pediatric dosing regimens for dystonia, except for the maximum dose of carbidopa/levodopa. Published regimens use maximum doses of levodopa as high as 400 mg/day for the treatment of dystonia in CP^16^, but the most common maximum daily dose used by respondents was 100 mg/day. Noting that dystonia-specific dosing information is lacking for baclofen^17–19^, clonazepam^20^, and diazepam^18,21^, respondents used dosing regimens in line with those published for spasticity or seizure management.

**Table 5.**
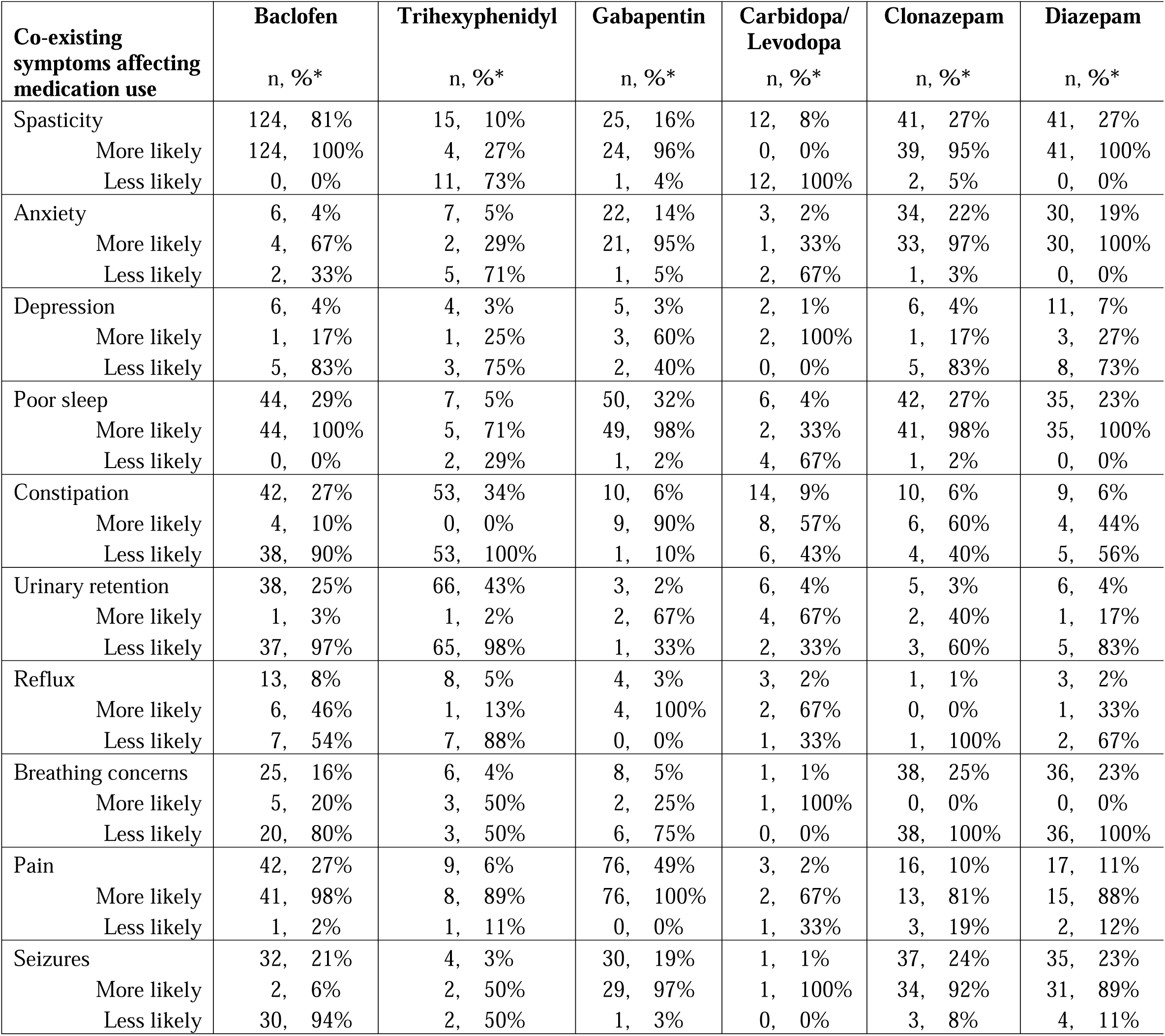
Co-existing symptoms affecting the choice to prescribe a medication. *% -Percentages are indicated in two ways: 1) Out of the total N of 154 (when giving the % of respondents stating that a specific co-existing symptom affects their choice to prescribe a medication), or 2) Out of the number of respondents stating that a given co-existing symptom affects their choice to prescribe a medication (when giving the % of respondents stating that they are more or less likely to prescribe a given medication in the presence of a given co-existing symptom). E.g. Spasticity affects the choice to prescribe baclofen for 124/154 respondents (81%). Of those respondents, 100% (124/124) stated that the presence of co-existing spasticity would make them more likely to prescribe baclofen.

**Table 6.** Respondent’s preferred dosing for enteral medications used to treat dystonia in CP. Respondents were given a choice between providing mg/day dosing and mg/kg/day dosing with the assumption that mg/kg/day dosing would be the preferred dosing paradigm for younger children while mg/day dosing might be preferred for adolescents and young adults. *Dosing used by respondents largely paralleled published dosing regimens except for the maximum dose of carbidopa/levodopa, where published dosing regimens use maximum doses as high as 400 mg/day.^16^ Note: maximum prescribed mg/day doses in the table may exceed safe maximal doses for children.

|  | **Baclofen** | **Trihexyphenidyl** | **Gabapentin** | **Carbidopa/**<br>**Levodopa** | **Clonazepam** | **Diazepam** |
| --- | --- | --- | --- | --- | --- | --- |
| **Starting dose**<br>**(median)** |  |  |  |  |  |  |
| mg/day | 5 | 1 | 100 | 25 | 0.25 | 1.75 |
| mg/kg/day | 0.5 | 0.1 | 10 | 1 | 0.02 | 0.12 |
| frequency/day | 2 | 2 | 3 | 2 | 2 | 3 |
| **Starting dose**<br>**(mode)** |  |  |  |  |  |  |
| mg/day | 5 | 1 | 300 | 25 | 0.25 | 1 |
| mg/kg/day | 0.5 | 0.1 | 10 | 1 | 0.01 | 0.1 |
| frequency/day | 3 | 2 | 3 | 3 | 2 | 3 |
| **Maximum dose**<br>**(median)** |  |  |  |  |  |  |
| mg/day | 80 | 15 | 2400 | 150* | 3 | 15 |
| mg/kg/day | 2 | 0.75 | 42.5 | 10 | 0.2 | 0.8 |
| frequency/day | 3 | 3 | 3 | 3 | 3 | 4 |
| **Maximum dose**<br>**(mode)** |  |  |  |  |  |  |
| mg/day | 80 | 60 | 3600 | 100* | 2 | 15 |
| mg/kg/day | 2 | 0.75 | 50 | 10 | 0.2 | 0.8 |
| frequency/day | 3 | 3 | 3 | 3 | 3 | 4 |

The co-existing symptoms most frequently affecting medication choice were spasticity, pain, urinary retention, and constipation. Eighty-one percent (124/154) of respondents would be more likely to prescribe baclofen in the setting of co-existing spasticity. Forty-nine percent (76/154) of respondents would be more likely to prescribe gabapentin in the setting of co-existing pain. Forty-two percent (65/154) and 34% (53/154) of respondents would be less likely to prescribe trihexyphenidyl in the setting of co-existing urinary retention or constipation, respectively. Noting that the above co-existing symptoms were explicitly queried as a part of the survey, the single most common co-existing symptom written-in by respondents was sialorrhea: 10% of respondents (15/154) would be more likely to prescribe trihexyphenidyl in the setting of sialorrhea. A summary of these preferred indications and dosing regimens is provided in Table 7.

**Table 7.** Summary of the indications and dosing cited by respondents for their six most frequently used enteral medications to treat dystonia in CP. *Most commonly used medications are indicated based on the relative frequency with which respondents stated they used each medication as 1^st^ line, 2^nd^ or 3^rd^ line, or 3^rd^ or 4^th^ line treatment choices (Table 4). **Potential indications and contraindications refer to co-existing symptoms that met two criteria (Table 5): 1) at least 20% of respondents felt the co-existing symptom would affect their choice to prescribe a medication, and 2) More than 90% of those respondents stated that the co-existing symptom would make them more likely to prescribe the medication (indication) or less likely to prescribe the medication (contraindication). #Starting and maximum doses refer to the most common dosing regimens used by the respondents (Table 6). Respondents were given a choice between providing mg/day dosing and mg/kg/day dosing with the assumption that mg/kg/day dosing would be the preferred dosing paradigm for younger children while mg/day dosing might be preferred for adolescents and young adults. ##Dosing used by respondents largely paralleled published dosing regimens except for the maximum dose of carbidopa/levodopa, where published dosing regimens use maximum doses as high as 400 mg/day.^16^ Note: maximum prescribed mg/day doses in the table may exceed safe maximal doses for children by weight. BID – twice a day, TID – three times a day, QID – four times a day

|  | **Baclofen** | **Trihexyphenidyl** | **Gabapentin** | **Carbidopa/**<br>**Levodopa** | **Clonazepam** | **Diazepam** |
| --- | --- | --- | --- | --- | --- | --- |
| **Most commonly used\*** | 1<sup>st</sup> line | 2<sup>nd</sup> or 3<sup>rd</sup> line | 2<sup>nd</sup> or 3<sup>rd</sup> line | 3<sup>rd</sup> or 4<sup>th</sup> line | 2<sup>nd</sup> or 3<sup>rd</sup> line | 3<sup>rd</sup> or 4<sup>th</sup> line |
| **Potential indications\*\*** | Spasticity, poor sleep, pain | - | Poor sleep, pain | - | Spasticity, anxiety, poor sleep, seizures | Spasticity, poor sleep, seizures |
| **Potential contraindications\*\*** | Constipation, urinary retention, seizures | Constipation, urinary retention | - | - | Respiratory difficulties | Respiratory difficulties |
| **Starting dose#** |  |  |  |  |  |  |
| mg/day | 5 | 1 | 300 | 25 | 0.25 | 1 |
| mg/kg/day | 0.5 | 0.1 | 10 | 1 | 0.01 | 0.1 |
| dose frequency | TID | BID | TID | TID | BID | TID |
| **Max dose#** |  |  |  |  |  |  |
| mg/day | 80 | 60 | 3600 | 100## | 2 | 15 |
| mg/kg/day | 2 | 0.75 | 50 | 10 | 0.2 | 0.8 |
| dose frequency | TID | TID | TID | TID | TID | QID |

Half of respondents would be comfortable randomizing their patients to receive any of the six most commonly used medications as a part of a clinical trial comparing their effectiveness for treating dystonia in CP (79/154, 51%). A large minority (56/154, 36%) would be comfortable randomizing their patients to receive any of the 10 explicitly queried medications (baclofen, trihexyphenidyl, gabapentin, carbidopa/levodopa, clonazepam, diazepam, clonidine, tetrabenazine, clobazam, and cannabidiol).

Of the respondents who provided additional comments at the end of the survey, 39% (14/36) noted the value of establishing current treatment practices to inform a clinical trial comparing the efficacy of these medications:

> *“Much more information is needed on treatment, efficacy and standard practices for treating dystonia. I appreciate the time spent on this study.”*

> *“Important work. There is need for clarity here.”*

> *“I would be comfortable randomizing to almost anything with guidance and support.”*

## Discussion

Dystonia in CP is a common condition lacking clear data to support enteral pharmacologic treatment. The current AACPDM treatment guideline, based largely on expert opinion, suggests the use of baclofen first line, trihexyphenidyl second line, gabapentin in people with pain, and clonidine in people with poor sleep.^10^ We demonstrate that physicians who treat dystonia in people with CP in the US and Canada prioritize functional impact and whether the dystonia is generalized when deciding whether to prescribe medications to treat dystonia in CP. They most commonly use a subset of six medications: baclofen, trihexyphenidyl, gabapentin, carbidopa/levodopa, clonazepam, and diazepam. Respondents prefer baclofen in people with co-existing spasticity, gabapentin for those with co-existing pain, carbidopa/levodopa for those with genetic or idiopathic dystonia etiologies, and they avoid trihexyphenidyl in people with constipation or urinary retention. They largely follow published dosing regimens for dystonia,^13–16^ when available, but otherwise follow published regimens for other symptoms that are often present in people with CP (e.g. spasticity and seizures).^17–21^ Though there appears to be some consensus on the preferred first-line medication (baclofen), there is no clear consensus on the choice of second or third-line medications. Finally, though respondents noted preferences for medications, indications, and dosing, they were still largely comfortable randomizing their patients to receive any of the commonly used medications used to treat dystonia in CP, with some noting the clear need for such a trial.

The lack of evidence supporting these treatment practices remains glaring.^8^ Though our results show that baclofen is the most commonly used first line medication to treat dystonia in CP, there are no controlled or prospective studies supporting baclofen’s use for this purpose.^8^ To provide the CP population with an evidence-based treatment paradigm for dystonia, it is necessary to do a placebo-controlled trial assessing the efficacy of enteral baclofen as a first line treatment of dystonia in CP. Furthermore, the lack of consensus regarding second- or third-line treatments necessitates a clinical trial comparing the effectiveness of these medications directly.

Limitations of this study center around the survey design. There was just over a 50% response rate, which may limit generalizability of the survey. Despite surveying a broad population of physicians who treat people with CP across two professional organizations, respondents were largely limited to 2 subspecialties: neurology and physiatry. This may, however, accurately reflect the physician populations most commonly prescribing these medications. We assessed only what physicians said were their prescribing practices, not their actual prescribing practices.

This work would be complemented by a study examining a large electronic medical record (EMR) database. However, it is important to note that an EMR-based study would indicate which medications are most commonly prescribed to people with ICD-10 diagnoses of dystonia and CP but would not be reliable in generating the preferred dosing regimens and indications for these medications, including whether a given medication was truly prescribed for dystonia or to treat another co-existing symptom.

In conclusion, this study summarizes the current indications and dosing for the six most commonly used medications to treat dystonia in people with CP according to physicians who treat this population in the US and Canada (Table 7). The survey also demonstrates physician support for a trial comparing the effectiveness of pharmacologic treatments for dystonia in CP. This data may serve as a rough guide for trainees or other physicians interested in treating dystonia in this population and may also inform a rational dosing guide for assessing the comparative effectiveness of these medications in a clinical trial.

---

**Supplementary Table 1.** Medical specialties of survey respondents and non-respondents.

|  | Respondents | Non-respondents |
| --- | --- | --- |
| Specialty | n, % | n, % |
| Physiatry | 83, 35% | 72, 30% |
| Neurology/Neurodevelopmental Disabilities | 74, 31% | 82, 34% |
| Orthopedics | 38, 16% | 39, 16% |
| Developmental Pediatrics | 13, 5% | 18, 8% |
| Pediatrics - Other | 5, 2% | 15, 6% |
| Other | 5, 2% | 13, 5% |
| Not currently in clinical practice | 20, 8% | unknown -- |
| Unspecified | 2, 1% | 0, 0% |
| TOTAL | 240, 100% | 239, 100% |

**Supplementary Table 2.** Medications used to treat dystonia in CP and frequency of use: comparison by specialty and by number of years in practice. Medications are listed by descending frequency of use based on the results from all respondents. *Other – only 2 respondents wrote in medications they used that were different from the 10 medications explicitly queried: amantadine and benztropine.

|  | **Overall**<br>**(n=154)** | **Neurology**<br>**(n=56)** | **Physiatry**<br>**(n=74)** | **<5 yrs in**<br>**practice**<br>**(n=37)** | **6-15 yrs in**<br>**practice**<br>**(n=52)** | **>15 yrs in**<br>**practice**<br>**(n=66)** |
| --- | --- | --- | --- | --- | --- | --- |
| **Medications** | n, % | n, % | n, % | n, % | n, % | n, % |
| Baclofen | 147, 95% | 52, 93% | 72, 97% | 33, 89% | 51, 98% | 64, 97% |
| Trihexyphenidyl | 122, 79% | 44, 79% | 65, 88% | 26, 70% | 43, 83% | 53, 80% |
| Gabapentin | 103, 67% | 34, 61% | 52, 70% | 28, 76% | 37, 71% | 38, 58% |
| Clonazepam | 85, 55% | 32, 57% | 43, 58% | 23, 62% | 32, 62% | 31, 47% |
| Carbidopa/Levodopa | 84, 55% | 32, 57% | 44, 59% | 15, 41% | 29, 56% | 41, 62% |
| Diazepam | 83, 54% | 26, 46% | 44, 59% | 23, 62% | 26, 50% | 34, 52% |
| Clonidine | 30, 19% | 16, 29% | 8, 11% | 11, 30% | 6, 12% | 13, 20% |
| Tetrabenazine | 27, 18% | 14, 25% | 11, 15% | 2, 5% | 11, 21% | 14, 21% |
| Clobazam | 11, 7% | 9, 16% | 1, 1% | 4, 11% | 4, 8% | 3, 5% |
| Cannabidiol | 7, 5% | 2, 4% | 2, 3% | 0, 0% | 2, 4% | 5, 8% |
| Other* | 2, 1% | 0, 0% | 2, 3% | 0, 0% | 1, 2% | 1, 2% |

