## Supplementary Methods for "Physician Approaches to the Pharmacologic Treatment of Dystonia in Cerebral Palsy"

### Medications used to treat dystonia in people with CP: Survey of current practices

We want to understand how you approach prescribing medications to treat dystonia in people with cerebral palsy. Your responses will be anonymous.

If you DO NOT currently provide medical care to people with cerebral palsy:
Please answer the first question, which will prompt you to exit the survey. We want to make sure we have accounted for you and avoid sending you survey completion reminders.

If you DO NOT prescribe medications to treat dystonia in people with cerebral palsy:
Please proceed through the first few questions of the survey. There will be a stop point prompting you to exit the survey. We anticipate that the portion of the survey applicable to you will take 1-2 minutes to complete.

If you do currently prescribe medications to treat dystona in people with cerebral palsy:
We anticipate that this survey will take you 15-20 MINUTES TO COMPLETE. Please be as thorough as possible with your responses. We thank you greatly for your time and feel strongly that your responses can guide a comprehensive approach to dystonia treatment in people with cerebral palsy.

You will be emailed your survey responses when you complete the survey, which you may find to be a useful compendium of your approach to treating dystonia in people with cerebral palsy. You may choose to share this compendium with your trainees or colleagues.

You can save your responses and return to the survey at a later time (scroll to the bottom, click "Save & Return Later." RedCap will send you an email with your specific survey link).

#### PROVIDER DEMOGRAPHICS

Do you currently provide medical care to people with cerebral palsy?

- ◯ Yes
- ◯ No (will end the survey)

What percentage of your patients have cerebral palsy? Please take your best guess.

- ◯ < 5
- ◯ 5-25
- ◯ 26-50
- ◯ 51-75
- ◯ >75

What percentage of your patients with cerebral palsy have dystonia?
Please take your best guess.

- ◯ < 5
- ◯ 5-25
- ◯ 26-50
- ◯ 51-75
- ◯ >75

Do you primarily treat adults, children, or both?

- ◯ Adults
- ◯ Children
- ◯ Both

Do you primarily practice in an academic center, private practice center, or a mix of the two?

- ◯ Academic
- ◯ Private practice
- ◯ Mix of academic and private practice

In what country do you practice?

- ◯ United States
- ◯ Canada
- ◯ Other

Please name the country: ______________________

| | |
|---|---|
| What is your medical speciality? | ◯ Physiatry<br>◯ Neurology<br>◯ Developmental Pediatrics<br>◯ Orthopedics<br>◯ Neurosurgery<br>◯ Other (please describe) |
| Please describe specialty: | __________________________________ |
| Did you do fellowship training after residency?<br>If yes, please describe. | ◯ Yes (please describe) ______<br>◯ No |
| How long have you been practicing independently? | ◯ I am still in training (resident/fellow)<br>◯ 0-5 years<br>◯ 6-10 years<br>◯ 11-15 years<br>◯ >15 years |

#### DYSTONIA ASSESSMENT

How do you typically assess for dystonia in your clinical practice? Check all that apply.

- ☐ I typically do NOT assess for dystonia in my clinical practice.
- ☐ History
- ☐ Physical exam/neurological exam
- ☐ Post-clinic review of their motor exam video ______
- ☐ A standardized assessment tool/scale ______
- ☐ Other way to assess dystonia (please describe) ______

#### MEDICAL FEATURES AFFECTING PRESCRIBING PRACTICES

| | |
|---|---|
| Do you currently prescribe medications to people with cerebral palsy as part of your clinical practice? | ◯ Yes<br>◯ No (will end the survey) |
| Do you currently prescribe medications to treat dystonia in people with cerebral palsy as part of your clinical practice? | ◯ Yes<br>◯ No (will end the survey) |
| Which medical features, if any, would help you determine WHETHER OR NOT TO PRESCRIBE MEDICATIONS AT ALL for dystonia treatment in a person with cerebral palsy?<br><br>Select all that apply and please explain. | ☐ Age ______<br>☐ Severity (based on my assessment in the clinic) ______<br>☐ Focal vs. generalized dystonia ______<br>☐ Body regions involved (e.g. dystonia in the arms vs. in the legs) ______<br>☐ Interference / functional impact (i.e. whether a person's dystonia prevents their ability to do a task that they deem is important to them, causes significant pain, interferes with sleep, or creates challenges associated with ease of caregiving) ______<br>☐ Gross motor functional status ______<br>☐ Etiology of cerebral palsy ______<br>☐ Medical complexity (e.g. need for G-tube, tracheostomy, or other specialty-based medical care) ______<br>☐ Prevention of secondary musculoskeletal concerns ______<br>☐ Perioperative tone management ______<br>☐ Other factors (please list and describe) ______ |
| If you feel a person with cerebral palsy meets your criteria for prescribing a dystonia medication, which medical features, if any, would affect your CHOICE OF MEDICATION(S)?<br><br>Select all that apply and please explain. | ☐ Age ______<br>☐ Severity (based on my assessment in the clinic) ______<br>☐ Focal vs. generalized dystonia ______<br>☐ Body regions involved (e.g. dystonia in the arms vs. in the legs) ______<br>☐ Interference / functional impact (i.e. whether a person's dystonia prevents their ability to do a task that they deem is important to them, causes significant pain, interferes with sleep, or creates challenges associated with ease of caregiving) ______<br>☐ Gross motor functional status ______<br>☐ Etiology of cerebral palsy ______<br>☐ Medical complexity (e.g. need for G-tube, tracheostomy, or other specialty-based medical care) ______<br>☐ Prevention of secondary musculoskeletal concerns ______<br>☐ Perioperative tone management ______<br>☐ Other factors (please list and describe) ______ |

#### MEDICATION EFFICACY

How do you typically assess the efficacy of the medications you use to treat dystonia in people with cerebral palsy? Select all that apply

- ☐ Asking the person with CP and/or their caregivers if they feel there has been an improvement
- ☐ Establishing a shared treatment goal BEFORE starting the medication and asking the person with CP and/or their caregivers about progress toward obtaining that goal
- ☐ Serial assessments of dystonia severity in the clinic based on your physical exam
- ☐ Serial assessments of dystonia severity using a validated scale ______
- ☐ Determining whether the burden of undesirable treatment side effects outweighs any benefits seen
- ☐ Other (please describe) ______

If a current medication at its goal/maximum dose is not efficacious for the treatment of dystonia in a person with cerebral palsy, what do you typically do next?

- ◯ Wean off the current medication first before adding a new medication
- ◯ Add a new medication without first weaning off the current medication
- ◯ Simultaneously wean the current medication while adding and up-titrating the new medication
- ◯ Other (please explain) ______

#### MOST COMMONLY USED MEDICATIONS

What are the medications you most commonly use to treat dystonia in people with cerebral palsy (including both initial treatment and subsequent treatment)? Pick your top 5

MEDICATION KEY
BAC - Baclofen, CBD - Cannabidiol, LDOPA - Carbidopa/Levodopa, CLB- Clobazam, CZP - Clonazepam, DZP - Diazepam, GBP- Gabapentin, TBZ - Tetrabenazine, THP - Trihexyphenidyl

| | BAC | CBD | LDOPA | CLB | Clonidine | CZP | DZP | GBP | TBZ | THP | Other |
|---|---|---|---|---|---|---|---|---|---|---|---|
| 1st most commonly used | ○ | ○ | ○ | ○ | ○ | ○ | ○ | ○ | ○ | ○ | ○ |
| 2nd most commonly used | ○ | ○ | ○ | ○ | ○ | ○ | ○ | ○ | ○ | ○ | ○ |
| 3rd most commonly used | ○ | ○ | ○ | ○ | ○ | ○ | ○ | ○ | ○ | ○ | ○ |
| 4th most commonly used | ○ | ○ | ○ | ○ | ○ | ○ | ○ | ○ | ○ | ○ | ○ |
| 5th most commonly used | ○ | ○ | ○ | ○ | ○ | ○ | ○ | ○ | ○ | ○ | ○ |

Please list generic name of "other" medication:

__________________________________

__________________________________

__________________________________

__________________________________

__________________________________

__________________________________

**CO-EXISTING SYMPTOMS and MEDICATION DOSING**

**Please enter dosing numerically in mg/day OR mg/kg/day, based on your preference. Include how often you typically dose each medicaton per day (noting that this may not be universal across your clinical practice). You may provide rationale for your responses or indicate other factors you take into account in the "Comments" column.**

1st most commonly used medication:
[first_choice_medname]

| | |
|---|---|
| Please select any co-existing symptom(s) that would affect the likelihood of you prescribing [first_choice_medname].<br><br>Then, indicate whether it makes you MORE or LESS likely to prescribe [first_choice_medname].*<br><br>Leave checkbox blank if N/A. | ☐ Spasticity ______<br>☐ Anxiety ______<br>☐ Depression ______<br>☐ Poor sleep ______<br>☐ Constipation ______<br>☐ Urinary retention ______<br>☐ Reflux ______<br>☐ Breathing concerns ______<br>☐ Pain ______<br>☐ Seizures ______<br>☐ Other (please enter symptom) ______ ______ |

*Please describe how you dose [first_choice_medname].

mg/day OR mg/kg/day # of daily doses Comments
Starting dose ______ OR ______ ______ ______
Maximum dose ______ OR ______ ______ ______
*enter dosing in mg/day OR mg/kg/day*

2nd most commonly used medication:
[second_choice_medname]

| | |
|---|---|
| Please select any co-existing symptom that would affect the likelihood of you prescribing [second_choice_medname].<br><br>Then, indicate whether it makes you MORE or LESS likely to prescribe [second_choice_medname].*<br><br>Leave checkbox blank if N/A. | ☐ Spasticity ______<br>☐ Anxiety ______<br>☐ Depression ______<br>☐ Poor sleep ______<br>☐ Constipation ______<br>☐ Urinary retention ______<br>☐ Reflux ______<br>☐ Breathing concerns ______<br>☐ Pain ______<br>☐ Seizures ______<br>☐ Other (please enter symptom) ______ ______ |

*Please describe how you dose [second_choice_medname].

mg/day OR mg/kg/day # of daily doses Comments
Starting dose ______ OR ______ ______ ______
Maximum dose ______ OR ______ ______ ______
*enter dosing in mg/day OR mg/kg/day*

3rd most commonly used medication:
[third_choice_medname]

Please select any co-existing symptom that would affect the likelihood of you prescribing [third_choice_medname].

Then, indicate whether it makes you MORE or LESS likely to prescribe [third_choice_medname].*

Leave checkbox blank if N/A.

- ☐ Spasticity ______
- ☐ Anxiety ______
- ☐ Depression ______
- ☐ Poor sleep ______
- ☐ Constipation ______
- ☐ Urinary retention ______
- ☐ Reflux ______
- ☐ Breathing concerns ______
- ☐ Pain ______
- ☐ Seizures ______
- ☐ Other (please enter symptom) ______ ______

*Please describe how you dose [third_choice_medname].

| | mg/day | OR | mg/kg/day | # of daily doses | Comments |
|---|---|---|---|---|---|
| Starting dose | ______ | OR | ______ | ______ | ______ |
| Maximum dose | ______ | OR | ______ | ______ | ______ |

*enter dosing in mg/day OR mg/kg/day*

4th most commonly used medication:
[fourth_choice_medname]

Please select any co-existing symptom that would affect the likelihood of you prescribing [fourth_choice_medname].

Then, indicate whether it makes you MORE or LESS likely to prescribe [fourth_choice_medname].*

Leave checkbox blank if N/A.

- ☐ Spasticity ______
- ☐ Anxiety ______
- ☐ Depression ______
- ☐ Poor sleep ______
- ☐ Constipation ______
- ☐ Urinary retention ______
- ☐ Reflux ______
- ☐ Breathing concerns ______
- ☐ Pain ______
- ☐ Seizures ______
- ☐ Other (please enter symptom) ______ ______

*Please describe how you dose [fourth_choice_medname].

| | mg/day | OR | mg/kg/day | # of daily doses | Comments |
|---|---|---|---|---|---|
| Starting dose | ______ | OR | ______ | ______ | ______ |
| Maximum dose | ______ | OR | ______ | ______ | ______ |

*enter dosing in mg/day OR mg/kg/day*

5th most commonly used medication:
[fifth_choice_medname]

Please select any co-existing symptom that would affect the likelihood of you prescribing [fifth_choice_medname].

Then, indicate whether it makes you MORE or LESS likely to prescribe [fifth_choice_medname].*

Leave checkbox blank if N/A.

- ☐ Spasticity ______
- ☐ Anxiety ______
- ☐ Depression ______
- ☐ Poor sleep ______
- ☐ Constipation ______
- ☐ Urinary retention ______
- ☐ Reflux ______
- ☐ Breathing concerns ______
- ☐ Pain ______
- ☐ Seizures ______
- ☐ Other (please enter symptom) ______ ______

*Please describe how you dose [fifth_choice_medname].

| | mg/day | OR | mg/kg/day | # of daily doses | Comments |
|---|---|---|---|---|---|
| Starting dose | ______ | OR | ______ | ______ | ______ |
| Maximum dose | ______ | OR | ______ | ______ | ______ |

*enter dosing in mg/day OR mg/kg/day*

#### EQUIPOISE

Consider the following question in the context of a clinical trial assessing the efficacy of medications used for the initial treatment of dystonia in people with cerebral palsy. Participating investigators would be given dosing, uptitration, and monitoring instructions for the medications assessed.

What medication(s), if any, would you be comfortable randomizing your clinic patients to receiving for the initial treatment of dystonia in cerebral palsy?

Select all that apply.

- ☐ I would be comfortable randomizing my clinic patients to receive any of these medications
- ☐ Clonazepam
- ☐ Diazepam
- ☐ Clobazam
- ☐ Baclofen
- ☐ Trihexyphenidyl
- ☐ Clonidine
- ☐ Gabapentin
- ☐ Carbidopa/Levodopa
- ☐ Tetrabenazine
- ☐ Cannabidiol
- ☐ Other (list generic name) ______

#### FEEDBACK

Please describe any other thoughts or comments you would like to share.

________________________________________
